# Multi-chain Fudan-CCDC model for COVID-19 in Iran

**DOI:** 10.1101/2020.04.22.20075630

**Authors:** Hanshuang Pan, Nian Shao, Yue Yan, Xinyue Luo, Ali Ahmadi, Yasin Fadaei, Jin Cheng, Wenbin Chen

**Affiliations:** School of Mathematical Sciences, Fudan University, 220 Handan Road, Shanghai, 200433, China; Shanghai Key Laboratory for Contemporary Applied Mathematics, Fudan University, 220 Handan Road, Shanghai, 200433, China; School of Mathematics, Shanghai University of Finance and Economics, 777 Guoding Road, Shanghai, 200433, China; Modeling in Health Research Center School of Public Health, Shahrekord University of Medical Sciences, Shahrekord, 88155383, Iran

**Keywords:** COVID-19, Iran, multi-chain Fudan-CCDC model

## Abstract

**Background:** COVID-19 has been deeply affecting people’s lives all over the world. It is significant for prevention and control to model the evolution effectively and efficiently.

**Methods:** We first propose the multi-chain Fudan-CCDC model which is based on the original Fudan-CCDC model to describe the revival of COVID-19 in some countries. Multi-chains are considered as the superposition of distinctive single chains. Parameter identification is carried out by minimizing the penalty function.

**Results:** From results of numerical simulations, the multi-chain model performs well on data fitting and reasonably interprets the revival phenomena. The band of ±25% fluctuation of simulation results could contain most seemly unsteady increments.

**Conclusion:** The multi-chain model has better performance on data fitting in revival situations compared with the single-chain model. It is predicted by the three-chain model with data by Apr 21 that the epidemic curve of Iran would level off on round May 10, and the final cumulative confirmed cases would be around 88820. The upper bound of the 95% confidence interval would be around 96000.

## 1 Introduction

COVID-19 is a new pandemic disease and precise data on its epidemic spread are not available in Iran and in the world. Important questions in peoples mind are as follows: How many people have COVID-19 in Iran? What is the status of COVID-19 epidemic curve in Iran? How will the epidemic develop and when will it end? Those questions have been preliminarily investigated in terms of modeling and use the daily reports of definitive COVID-19 patients released by Iran Ministry of Health and Medical Education in [2].

In this paper, we apply the multi-chain Fudan-CCDC model to analyze COVID-19 situations in Iran and present our predictions. The Fudan-CCDC model has been proposed in [10, 9, 13] based on the TDD-NCP model [14, 4, 3, 6, 7, 12], where the time delay process was taken into account. It also introduced a series of new convolution kernels for the time delay terms by applying several time distributions acquired from an important paper [5] by CCDC (China Center for Disease Control and Prevention). Both the TDD-NCP model and the Fudan-CCDC model are single-chain models, and have been performed well in analyzing the evolution of COVID-19 in China, and its early stage of global transmission [11, 15]. When tracking with our models, we proposed a multi-chain model in [8] to model the epidemic trend and predict cases in Singapore and found that the multi-chain model had better performance on data fitting in revival situations of countries like Singapore.

## 2 Methods

We introduce two models in this part, the single-chain Fudan-CCDC model and the multi-chain Fudan-CCDC model, which are similar to [8]. The single-chain Fudan-CCDC model describes one chain of transmission. And the multi-chain Fudan-CCDC model assumes that there may more than one transmission chain in the country, due to new imported cases, the spread of the epidemic in different regions or other reasons.

### 2.1 The single-chain Fudan-CCDC model

As is mentioned in [11, 9, 10, 13, 15], our single-chain Fudan-CCDC model is as follows:

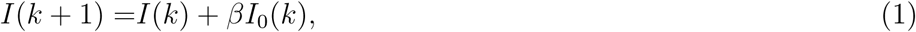

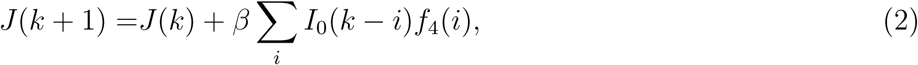

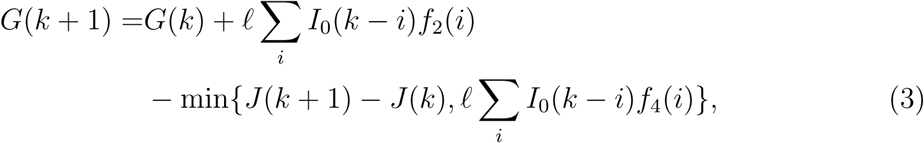

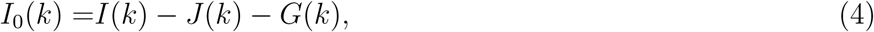

where *β* and *l* are the infection rate and the isolation rate respectively, which can be changed in time. *I*(*k*) and *J* (*k*) represent the cumulative infected people and the cumulative confirmed cases at day *k*, respectively, and *G*(*k*) is the instant (not cumulative) number of infected isolated not yet confirmed by the hospital. *I*_0_(*k*) is the number of people who are infected but not in quarantine or hospitalization. *f*_2_(*k*) and *f*_4_(*k*) are the transition probabilities from infection to illness onset, and from infection to hospitalization, respectively, which are extracted from [5] by CCDC. The kernels like *f*_2_(*k*) and *f*_4_(*k*) may be different in country. Note that the expression (3) is a little bit different from the one in [8] mainly considering the real situation.

The model can be used to fit the reported numbers of the cumulative confirmed cases and predict the evolution of epidemic, and the details can be found in [11, 9, 10, 13, 15].

### 2.2 The multi-chain Fudan-CCDC model

As is mentioned in [8], the multi-chain Fudan-CCDC model is the superposition of several single chains:

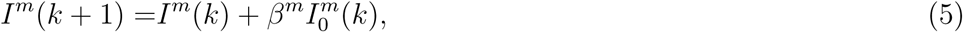

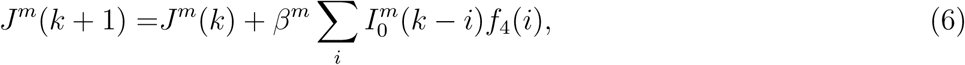

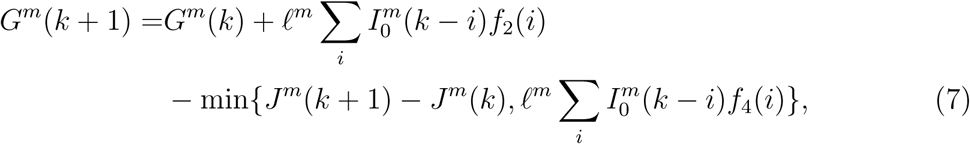

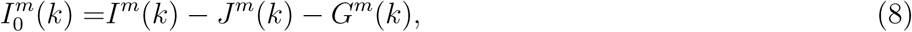

and we obtain the sum forms:

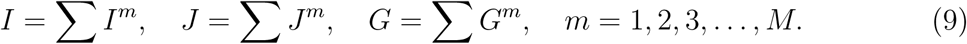

where *t*^*m*^ is the start time of the *m*-th source.

For both the multi-chain model, infection rate *β* and isolation rate *l* of the fisrt chain are obtained by fitting data before a specific time node. Considering that the isolation rate should be stable in a certain region, we suppose that it stays the same.

The optimization process is similar with the method used in [8], however we changed the objective function as follows:

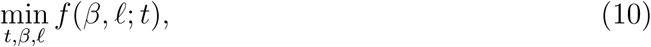

where *f* (*β, l* ; *t*) = ∥*J − data*∥_2_, and *data* means the public data of cumulative confirmed cases.

The unknown parameters were estimated by running the fminsearch, a MATLAB function. And we get the 95% confidence interval (CI) of our prediction by nlpredci.

## 3 Results

While tracking the Iran’s epidemic trend every day [2, 1], single chain model performed well at first. However, around March 17, we found that the daily increment did not drop down as the single-chain model had expected, while the data could be successfully identified by the two-chain model after March 19. As we mentioned in [8], when a sudden turn appears in the curve of reported confirmed cases, we have reason to suppose that there is a new chain. Around March 24 we can observe that in Iran’s epidemic trend.

Similarly, on April 10, there was a sudden increase in daily increment, which was twice the predicted value of the previous two-chain model. At the same time, we observed that the public data of total confirmed cases exceeded the maximum of the previous two-chain model, shown in Fig 1. That all aroused the suspicion whether the third chain was generated. We did reverse a new chain.

**Figure 1:**
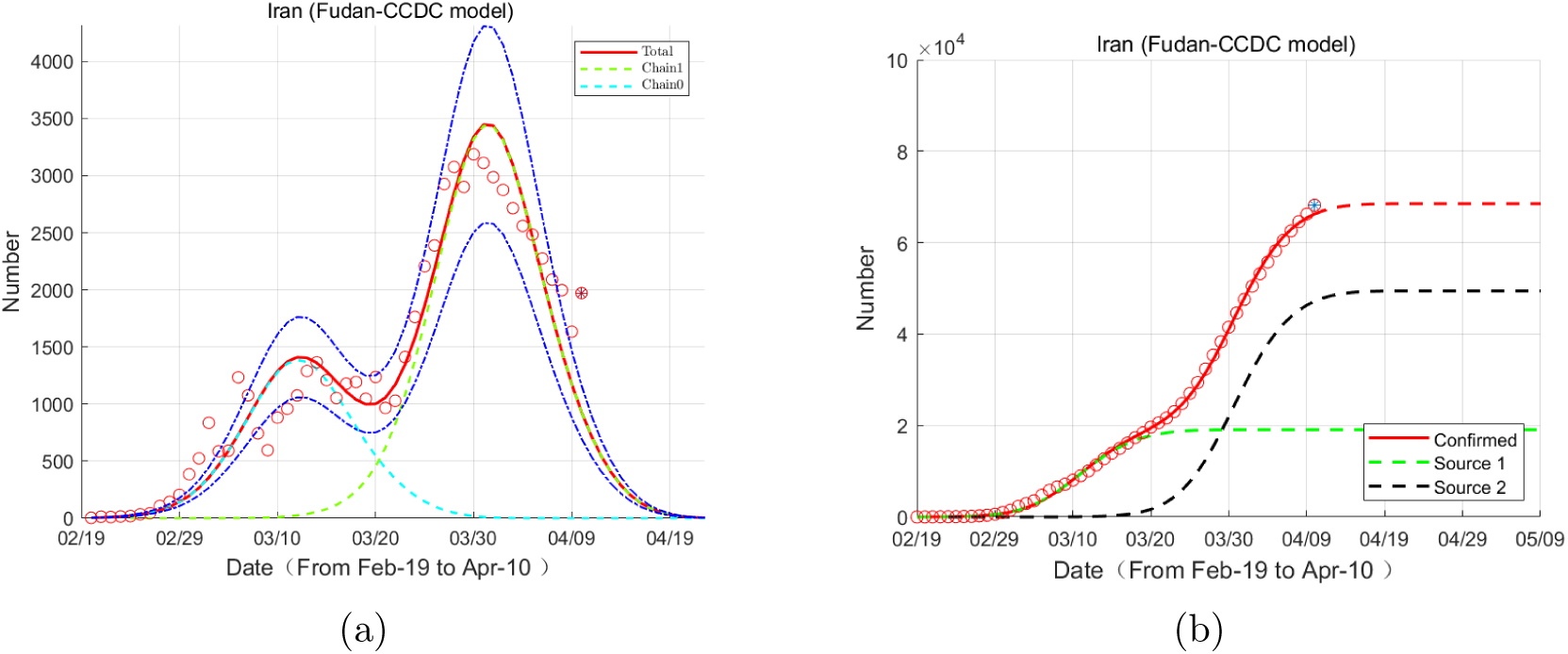
(a) Evolution of the increment of confirmed cases based on the two-chain Fudan-CCDC model. Red circles: data; Red lines: model predictions; (b) Evolution of number of cumulative confirmed cases based on the two-chain Fudan-CCDC model. Red circles: data; Red lines: model predictions; Green lines: first chain; Black lines: second chain. (b) The semi-log form of (a).

Figure 2 is our prediction of the Iran’s epidemic, based on the three-chain model, with data from Feb 19 to Apr 21. The results we obtained are as follows. In the absence of a new chain of transmission, the epidemic will eventually end on May 10, and the final number of the confirmed will be around 88820. The 95% confidence interval is [81740,95930].

**Figure 2:**
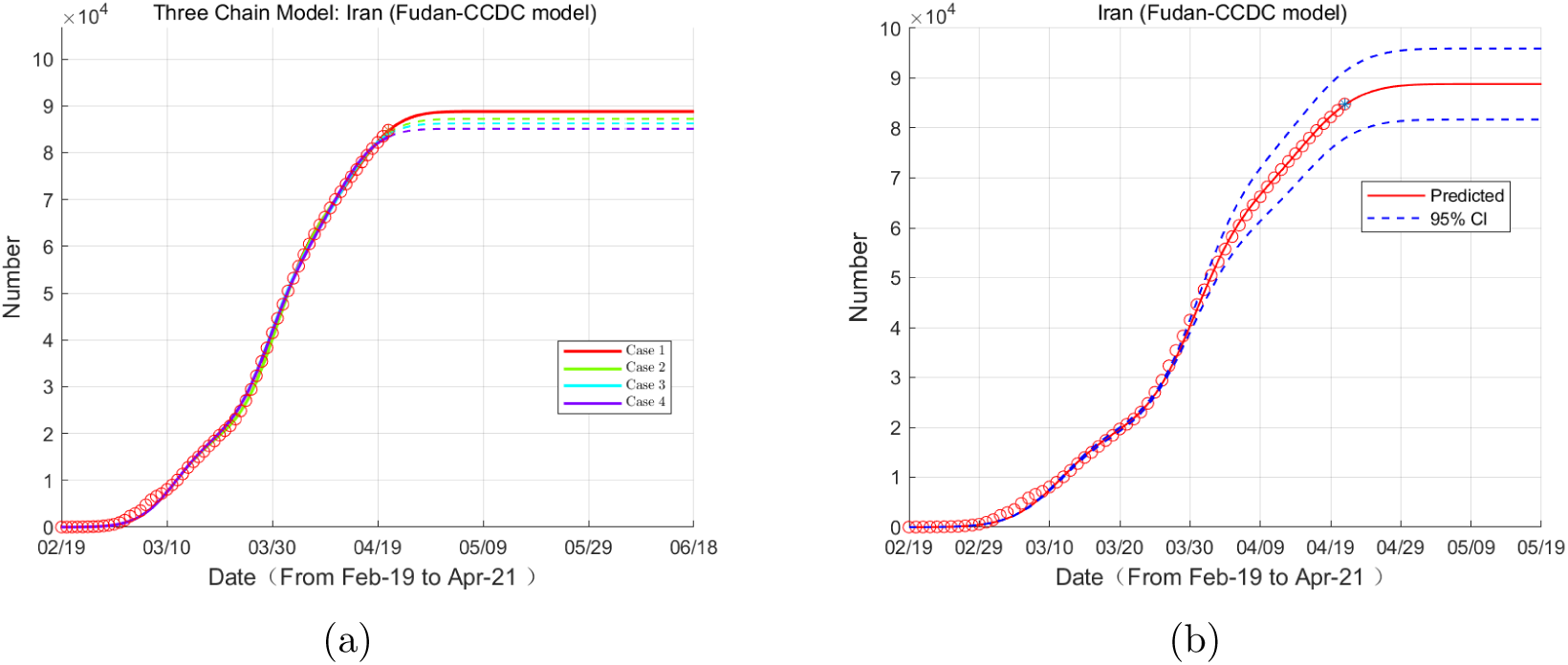
(a) Evolution of the number of cumulative confirmed cases based on the three-chain Fudan-CCDC model. Red circles: data; Red lines: model predictions; (b) Evolution of the number of cumulative confirmed cases based on the three-chain Fudan-CCDC model. Red circles: data; Red lines: model predictions; Blue dot line: 95% CI.

Figure 2(a) shows the evolutions of epidemic in Iran and its possible future trends.The scattered red circles are the data: the number of cumulative confirmed cases and its increment from Feb 19 to Apr 21. In Figure 2(a), we illustrate the four ‘most optimized’ fitting curves (in solid lines) for the data, and their predictions (in dotted lines) by the model, in the order of red, green, blue and purple, respectively. For the convenience to recognize, the ‘very most optimized’ fitting curve is drawn in a full solid red line, including its prediction. Details of the optimization methods are described in the section Method and [8].

In Figure 2(b), we show the 95% CI with blue lines, the scattered are still public data, the black line represents our predicted result.

Figure 3 is the result of our decomposition of the three chains of the previous epidemic, with data from Feb 19 to Apr 21. We can see the current stage has passed the peak of the latest chain. The shadow means the range of ±25% fluctuation from simulation results. From Figure 3(a), we can see the decomposition on total number of comfirmed cases.

**Figure 3:**
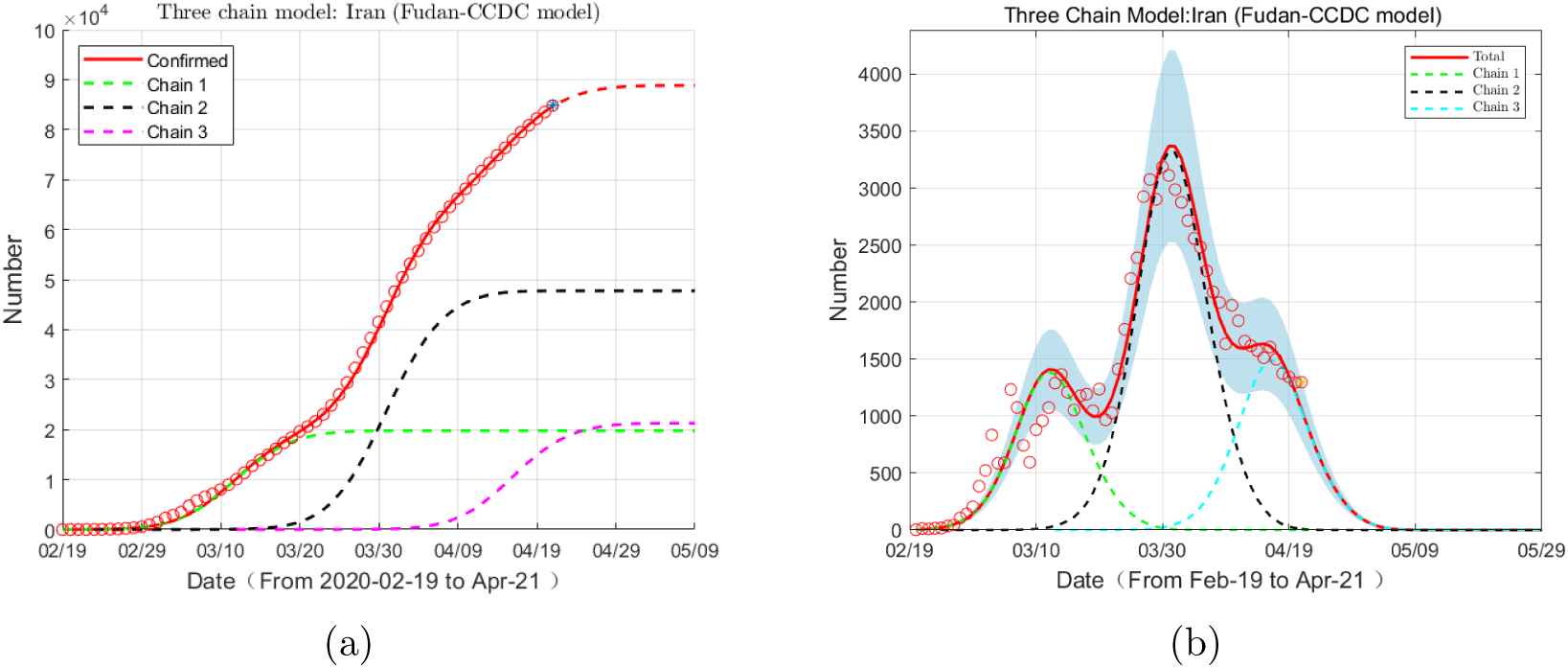
(a) Evolution of the number of cumulative confirmed cases based on the three-chain Fudan-CCDC model. Red circles: data; Red lines: model predictions; Green lines: first chain; Black lines: second chain; Blue lines: third chain. (b) Evolution of increment of confirmed cases based on the three-chain Fudan-CCDC model. Red circles: data; Red lines: model predictions; Green lines: first chain; Black lines: second chain; Blue lines: third chain.

From Figure 3(b), we can see the decomposition on increment instead.

Figure 4 shows our model has a good predictive effect when the third chain information is stable. The data of the hollow circles are used for prediction, and the filled circles are the true values afterwards. The red line is our forecast result.

**Figure 4:**
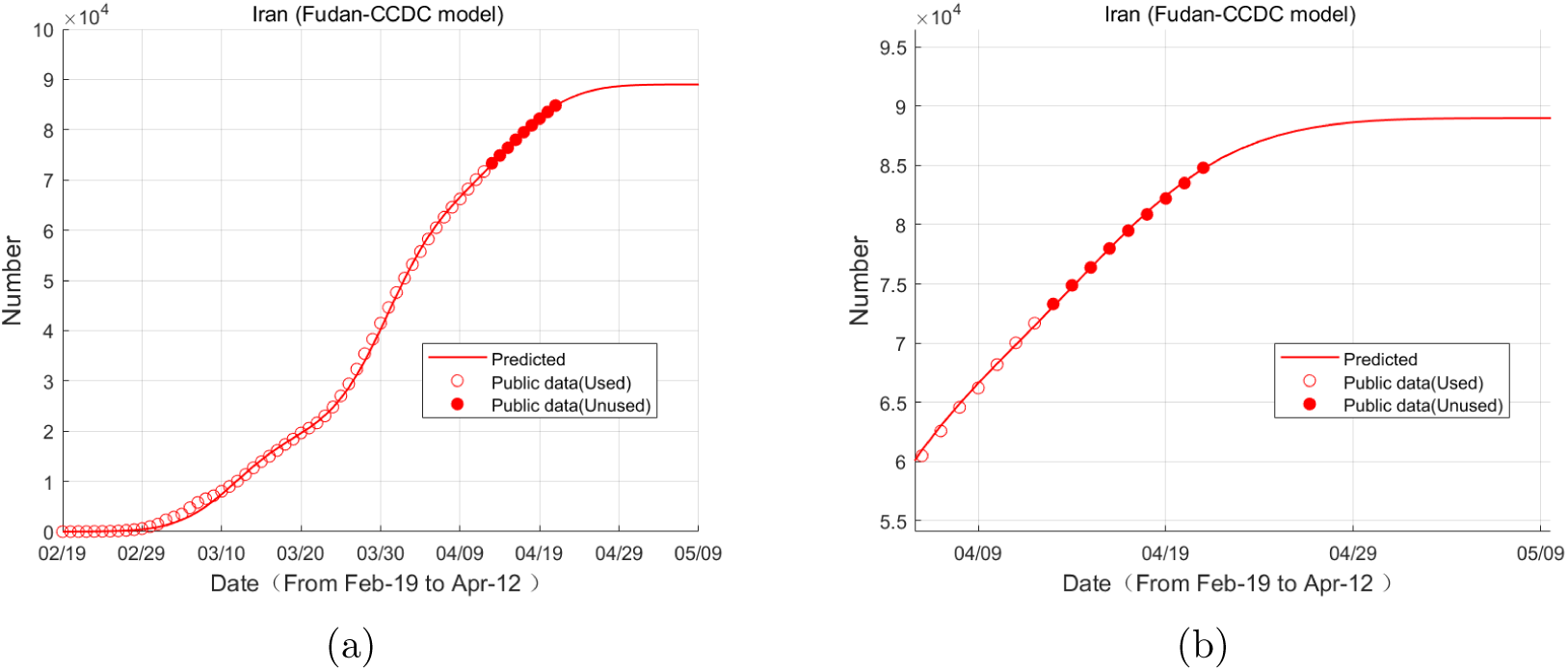
(a) Comparison of forecast results on April 12 and real data; (b) An enlarged version of (a).

### Parameter Table

In Table 1, the notations *t*^*j*^, *i*^*j*^, *r*^*j*^, *l* ^*j*^ represent *j*th chain’s parameters, the notation max is the predicted final infected number, and *t*_*end*_ represents the predicted end date.

**Table 1:** Parameters for three-chain model.

| Date | $t^0$ | $i^0$ | $r^0$ | $\ell^0$ | $t^1$ | $i^1$ | $r^1$ | $\ell^1$ | $t^2$ | $i^2$ | $r^2$ | $\ell^2$ | max | $t^{end}$ |
| --- | --- | --- | --- | --- | --- | --- | --- | --- | --- | --- | --- | --- | --- | --- |
| 2020/4/10 | 2020/2/13 | 1 | 0.91457 | 2.0368 | 2020/2/23 | 1 | 0.92202 | 2.0368 | 2020/3/12 | 1 | 0.90687 | 2.0368 | 77330 | 2020/5/5 |
| 2020/4/11 | 2020/2/13 | 1 | 0.91457 | 2.0368 | 2020/2/23 | 1 | 0.92198 | 2.0368 | 2020/3/12 | 1 | 0.91356 | 2.0368 | 85101 | 2020/5/7 |
| 2020/4/12 | 2020/2/13 | 1 | 0.91457 | 2.0368 | 2020/2/23 | 1 | 0.92197 | 2.0368 | 2020/3/12 | 1 | 0.91534 | 2.0368 | 88994 | 2020/5/8 |
| 2020/4/13 | 2020/2/13 | 1 | 0.91457 | 2.0368 | 2020/2/23 | 1 | 0.92196 | 2.0368 | 2020/3/12 | 1 | 0.9158 | 2.0368 | 90030 | 2020/5/10 |
| 2020/4/14 | 2020/2/13 | 1 | 0.91457 | 2.0368 | 2020/2/23 | 1 | 0.92196 | 2.0368 | 2020/3/12 | 1 | 0.91577 | 2.0368 | 89954 | 2020/5/10 |
| 2020/4/15 | 2020/2/13 | 1 | 0.91457 | 2.0368 | 2020/2/23 | 1 | 0.92196 | 2.0368 | 2020/3/12 | 1 | 0.91556 | 2.0368 | 89487 | 2020/5/10 |
| 2020/4/16 | 2020/2/13 | 1 | 0.91457 | 2.0368 | 2020/2/23 | 1 | 0.92197 | 2.0368 | 2020/3/12 | 1 | 0.91544 | 2.0368 | 89223 | 2020/5/10 |
| 2020/4/17 | 2020/2/13 | 1 | 0.91457 | 2.0368 | 2020/2/23 | 1 | 0.92197 | 2.0368 | 2020/3/12 | 1 | 0.91533 | 2.0368 | 89003 | 2020/5/10 |
| 2020/4/18 | 2020/2/13 | 1 | 0.91457 | 2.0368 | 2020/2/23 | 1 | 0.92197 | 2.0368 | 2020/3/12 | 1 | 0.91524 | 2.0368 | 88801 | 2020/5/10 |
| 2020/4/19 | 2020/2/13 | 1 | 0.91457 | 2.0368 | 2020/2/23 | 1 | 0.92198 | 2.0368 | 2020/3/12 | 1 | 0.91519 | 2.0368 | 88701 | 2020/5/10 |
| 2020/4/20 | 2020/2/13 | 1 | 0.91457 | 2.0368 | 2020/2/23 | 1 | 0.92198 | 2.0368 | 2020/3/12 | 1 | 0.91519 | 2.0368 | 88704 | 2020/5/10 |
| 2020/4/21 | 2020/2/13 | 1 | 0.91457 | 2.0368 | 2020/2/23 | 1 | 0.92197 | 2.0368 | 2020/3/12 | 1 | 0.91525 | 2.0368 | 88820 | 2020/5/10 |

## 4 Discussion

As so far, through simulations, we find that the evolution of the epidemic can be well explained by three-chain Fudan-CCDC model, although in real-life transmissions, the number of chains may be far more than three. It is very happy to see that the trend of the Iran’s epidemic is getting better and better. However, according to our model, it is not time to relax. But early detection and cutting off the source of transmission is still needed for preventing the spread of a new chain.

Observing the parameter table (see Table 1), we can see that each parameter is relatively stable. The final number of people predicted is not fluctuating too much, and the prediction of the end of the epidemic is stable after April 11. Since April 10 when we observed the new chain, the second chain has been performing more stable than the third chain. The change of results between April 10th and 11th was the largest during these days, mainly due to the large gap between the increments in the two consecutive days and the model prediction results, then the model was revised accordingly. It can be seen from Figure 3 that the prediction results of the model on April 12 have been able to accurately characterize the third chain, so that in the following week, the trend of the epidemic is in line with model prediction.

In the experiment, we can see that the newest chain is unstable at first, generally increasing. Compared with the results of April 10 and April 11, shown in Figure 5, the third chain had a jump. After April 11, the third chain tends to stabilize. There are two possible reasons. One is that as time goes by, the information exposed by the new chain increases, and after going out of the independent trend, the model is revised during optimization. Second, the propagation of the third chain itself is still unstable, and the intensity of the third chain itself is gradually increasing with the passage of time. If there are two reasons, it is of great significance to discover and block the transmission in time.

**Figure 5:**
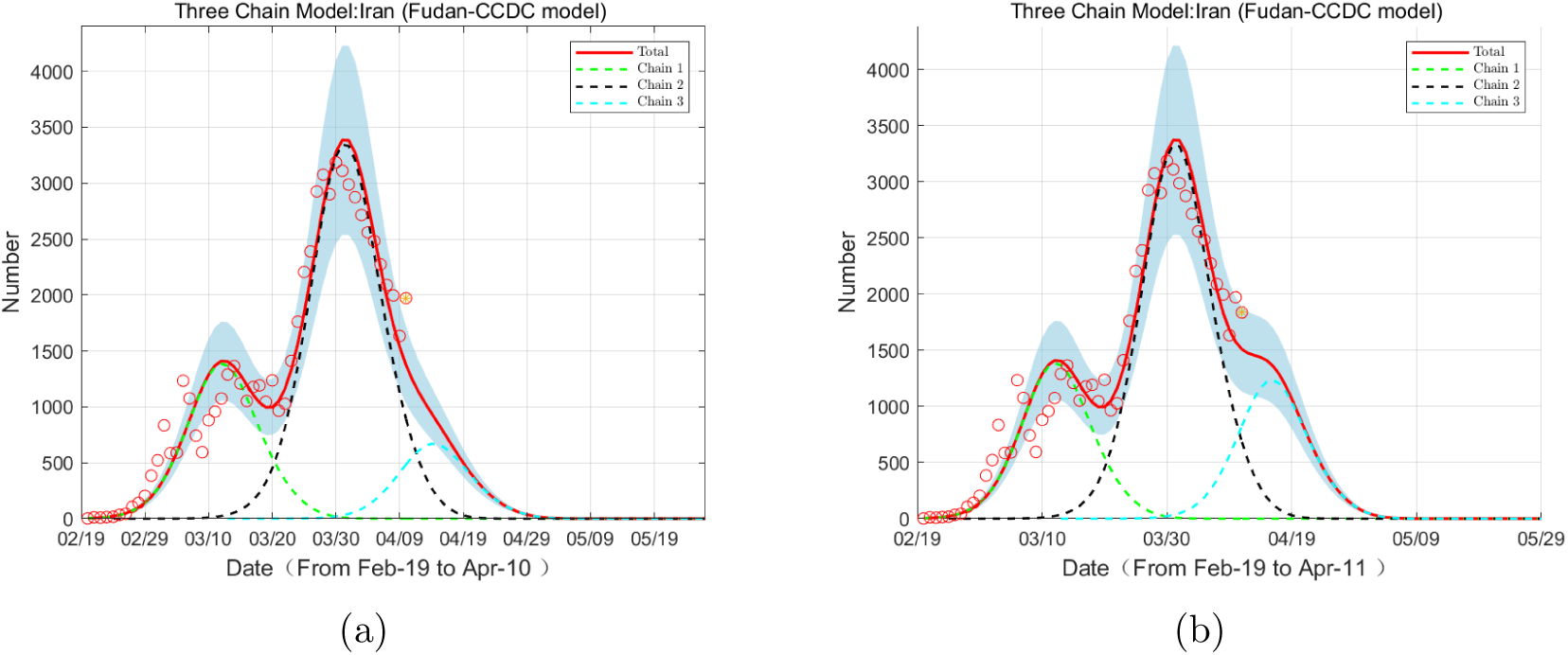
(a) Evolution of the number of increment confirmed cases based on the three-chain Fudan-CCDC model, with data from Feb 19 to Apr 10. Red circles: data; Red lines: model predictions; Green lines: first chain; Black lines: second chain; Blue lines: third chain. (b) Evolution of increment of confirmed cases based on the three-chain Fudan-CCDC model, with data from Feb 19 to Apr 11. Red circles: data; Red lines: model predictions; Green lines: first chain; Black lines: second chain; Blue lines: third chain.

## 5 Limitations

Its hard to verify the new chain at the very early stage. Only when the number of infections caused by the new chain reaches a certain relative size will it be discovered. This can be seen from the third chain of propagation from March 12, but it was not discovered until April 10. After all, the model only analyzes and interprets the data, and cannot determine the propagation in reality. All a good model can do is to discover the new chain of transmission as early as possible and evaluate its impact as accurately as possible, which will ultimately help in the judgment of epidemic decisions.

Model accuracy also depends on public data. For poor quality data, what the model can do is to propose better optimization methods and data processing methods. But the ultimate influence is ultimately limited. Good data will undoubtedly bring more accurate results. These may be improved in future work.

In our multi-chain model assumption, multiple propagation chains are independent and do not affect each other. But when multiple chains appear within a short period of time, do we need more delicate models to characterize? These questions will be improved in the next stage of work.

## 6 Conclusion

According to the data results as of April 21, the end for the Iran epidemic is about May 10, and the final number is about 88820.

At the same time, we need to pay close attention to the daily increment at this stage. When the daily increment does not drop down or even has a numerical rebound, it may indicate another chain, then the duration of the epidemic will be extended and the total number will also increase.

This article illustrates that it is of great significance to prevent the spread of new chains. To achieve this, people need to practice social distancing, detect and isolate the confected early.

## Data Availability

The data employed in this paper are acquired from the World Health Organization (https://www.who.int/emergencies/diseases/novel-coronavirus-2019/situation-reports). All the data can be accessed publicly. No other data are used in this paper.

https://www.who.in

## Acknowledgments

We are very grateful to the efforts of Cheng’s group members and the supports by School of Mathematical Sciences, Fudan University and School of Mathematics, Shanghai University of Finance and Economics. We thank Zhaojun Bai at UC Davis, Xiaoming Wang at South University of Science and Technology of China, Qiang Du at Columbia University, Long Chen at UC Irvine, Cheng Wang at University of Massachusetts at Dartmouth and Xingjie (Helen) Li at University of North Carolina at Charlotte. Wenbin Chen also thanks Wei Ge and Rongmin Li at Fudan University, Zhihua Shen at Fusion Fin Trade, Allen at Wind, Ning Liu at Winning Health Technology Group Company (winning.com.cn), Xinkang Cao and Jinwu Zhuo at Mathworks, and Weili Zhang. Ali and Yashi thank the Modeling in Health Research Center, School of Health, Deputy of Research and Technology, Shahrekord University of Medical Sciences. Furthermore, Ali and Yashi truly appreciate the Ministry of Health and Medical Education, especially the team of daily information reports of COVID-19 in Iran. All of us thank our families’ supports.

## Ethics approval and consent to participate

The ethical approval or individual consent was not applicable.

## Authors contributions

The algorithms are implemented by Hanshuang Pan, which are based on the single-chain model implemented by Nian Shao, and designed by Wenbin Chen. All authors conceived the study, carried out the analysis, discussed the results, drafted the first manuscript, critically read and revised the manuscript, and gave final approval for publication.

## Data and materials availability

The data employed in this paper are acquired from WIND (like Bloomberg), and the situation reports of the World Health Organization (url: https://www.who.int). All the data can be accessed publicly. No other data are used in this paper.

## References

[1] The National Committee on COVID-19 Epidemiology in Ministry of Health and Medical Education. Available from: http://corona.behdasht.gov.ir.

[2] A Ahmadi, Y Fadaei, M Shirani, and F Rahmani. Modeling and forecasting trend of COVID-19 epidemic in Iran until May 13, 2020. Med. J. Islam Repub. Iran, 34(1):183–195, 2020.

[3] Yu Chen, Jin Cheng, Yu Jiang, and Keji Liu. A time delay dynamic system with external source for the local outbreak of 2019-nCoV. Applicable Analysis, 2020. DOI: 10.1080/00036811.2020.1732357.

[4] Yu Chen, Jin Cheng, Yu Jiang, and Keji Liu. A time delay dynamical model for outbreak of 2019-nCoV and the parameter identification. J. Inverse Ill-posed Probl., 28(2):243–250, 2020.

[5] Qun Li, Xuhua Guan, Peng Wu, Xiaoye Wang, Lei Zhou, Yeqing Tong, Ruiqi Ren, Kathy SM Leung, Eric HY Lau, Jessica Y Wong, et al. Early transmission dynamics in Wuhan, China, of novel coronavirus–infected pneumonia. New Engl. J. Med., 382:1199–1207, 2020.

[6] Keji Liu, Yu Jiang, Yue Yan, and Wenbin Chen. A time delay dynamic model with external source and the basic reproductive number estimation for the outbreak of Novel Coronavirus Pneumonia. Control Theory and Appl. (in Chinese), 37(3):453–460, 2020.

[7] Xinyue Luo, Nian Shao, Jin Cheng, and Wenbin Chen. Modeling the trend of outbreak of COVID-19 in the Diamond Princess cruise ship based on a time-delay dynamic system. Math. Modeling Appl. (in Chinese), 9(1):15–22, 2020.

[8] Hanshuang Pan, Nian Shao, Yue Yan, Xinyue Luo, Shufen Wang, Ling Ye, Jin Cheng, and Wenbin Chen. Multi-chain Fudan-CCDC model for COVID-19 - a revisit to Singapore’s case. Preprint at https://www.medrxiv.org/content/early/2020/04/17/2020.04.13.20063792, 2020.

[9] Nian Shao, Yu Chen, Jin Cheng, and Wenbin Chen. Some novel statistical time delay dynamic model by statistics data from CCDC on Novel Coronavirus Pneumonia. Control Theory and Appl. (in Chinese), 37(4):697–704, 2020.

[10] Nian Shao, Jin Cheng, and Wenbin Chen. The reproductive number r_0_ of COVID-19 based on estimate of a statistical time delay dynamical system. Preprint at https://www.medrxiv.org/content/early/2020/02/20/2020.02.17.20023747.1, 2020.

[11] Nian Shao, Yan Xuan, Hanshuang Pan, Shufen Wang, Weijia Li, Yue Yan, Xingjie Li, Christopher Y. Shen, Xu Chen, Xinyue Luo, Yu Chen, Boxi Xu, Keji Liu, Min Zhong, Xiang Xu, Yu Jiang, Shuai Lu, Guanghong Ding, Jin Cheng, and Wenbin Chen. COVID-19 in Japan: what could happen in the future? Preprint at https://www.medrxiv.org/content/10.1101/2020.02.21.20026070v1.article-info., 2020.

[12] Nian Shao, Min Zhong, Jin Cheng, and Wenbin Chen. Modeling for COVID-19 and the prediction of the number of the infected based on fudan-ccdc. Math. Modeling Appl. (in Chinese), 9(1):29–32, 2020.

[13] Nian Shao, Min Zhong, Yue Yan, Pan Hanhuang, Jin Cheng, and Wenbin Chen. Dynamic models for CoVID-19 and data analysis. Math. Meth. Appl. Sci., 2020. DOI: 10.1002/mma.6345.

[14] Yue Yan, Yu Chen, Keji Liu, Xinyue Luo, Boxi Xu, Yu Jiang, and Jin Cheng. Modeling and prediction for the trend of outbreak of NCP based on a time-delay dynamic system. Sci. Sin. Math. (in Chinese), 50(3):385–392, 2020.

[15] Yue Yan, Hanshuang Pan, Nian Shao, Yan Xuan, Shufen Wang, Weijia Li, Xingjie Li, Christopher Y. Shen, Xu Chen, Xinyue Luo, Yu Chen, Boxi Xu, Keji Liu, Min Zhong, Xiang Xu, Yu Jiang, Shuai Lu, Guanghong Ding, Jin Cheng, and Wenbin Chen. COVID-19 in Singapore: another story of success. Int. J. Math. Industry, 2020. DOI: 10.1142/S266133522050001X.

